# Durability of the immune response to a third BNT162b2 dose; five months follow-up

**DOI:** 10.1101/2022.05.03.22274592

**Authors:** Mayan Gilboa, Gili Regev-Yochay, Michal Mandelboim, Victoria Indenbaum, Keren Asraf, Ronen Fluss, Sharon Amit, Ella Mendelson, Ram Doolman, Arnon Afek, Laurence S. Freedman, Yitshak Kreiss, Yaniv Lustig

## Abstract

**Background:** Two doses of the BNT162b2 vaccine yielded high effectiveness that wanes within several months. The third dose was effective in mounting a significant humoral and cellular immune response..

**Methods:** We followed BNT162b2-vaccinated health-care workers monthly for IgG and neutralizing antibody (NeutAb) titers. Avidity, T-cell activation and microneutralization of sera against different variants of concern (VOC) were assessed for a sub-cohort. Linear mixed models were used to compare the durability of the second and third doses, and to assess if Omicron breakthrough infections were associated with waning dynamics.

**Results:** Overall 3972 participants with a third dose were followed, the rate of waning of IgG and NeutAb was slower after the third (1.32%/day and 1.32%/day, respectively) compared to the second (2.26%/per day and 3.34%/day) dose. Live-neutralization of Omicron VOC was lower compared to previous strains and demonstrated similar waning from 111 (95%CI:75-166) to 26 (95%CI:16-42) within 4 months. Mean T cell activity decreased from 98±5.4 T cells/10^6^ PBMC to 59±9.3, within 3-5 months. Omicron breakthrough infections were associated with lower IgG peak (ratio of means 0.86 95%CI 0.80-0.91), and among participants over 65y with faster waning of both IgG and NeutAb (ratio of mean rates 1.40 95% CI 1.13-1.68 and 3.58 95% CI 1.92-6.67). No waining in IgG avidity was obsereved during 112 days after the 3^rd^ dose.

**Conclusion:** The third dose is more durable than the second dose, yet Omicron is relatively resistant to direct neutralization. The level of humoral response may predict breakthrough infections.

## Background

Two years after emerging, severe respiratory syndrome coronavirus 2 (SARS-CoV-2) pandemic, continues to take lives of thousands of people daily and to cause major economic, social and public health impairments[1]. Several vaccines have been shown to be highly efficacious and effective against infection and severe disease of SARS-CoV-2 and vaccine rollout is expanding worldwide[2–5]. While the best vaccine schedules are not yet determined, BNT162b2 was approved as a two-dose vaccine, with a 21 day interval between doses[6]. Several studies have demonstrated waning of immune responses and vaccine effectiveness of two doses within 6 months [7–9]. This lead the Israeli MOH, and later other countries to recommend a booster dose on July 29, 2021[10–12]. The third dose was shown to rapidly boost the immune response [13] and to induce high vaccine effectiveness [14] thus resulting in a superior immune response compared to the second dose[15]. Moreover, it was shown to be necessary for efficient neutralization of the Omicron variant of concern [16].While many countries are now recommending a third dose, the durability of a third dose effect is unknown.

Here we present the durability of the immune response to the third vaccine dose, within five months. We report the effect against various VOC’s and compare it to the immune response kinetics after the second dose. Last, we show how this immune response is associated with susceptibility to clinical infection five months after vaccination.

## Methods

### Ethical Statement

The protocol was approved by the Institutional Review Board of the Sheba Medical Centre (SMC) and written informed consent was obtained from all study participants.

### Study Design and Population

The Sheba Medical Centre, is the largest tertiary medical centre in Israel, with 1,600 beds and 15,480 HCW, including employees, students, volunteers, and retired personnel. HCW in SMC include 18% physicians, 27% nurses and nurse aids, 21% paramedical personnel and 34% administration and logistic employees.

The Sheba HCW COVID Cohort was initially established on March 2020, when 15,480 Sheba HCW were offered to join a sero-surveillance study. With the rollout of the COVID-19 vaccination campaign, monthly follow-up was offered to all participants. The recruitment and follow up of this cohort have been reported in detail previously [15,17–21]

Briefly, all HCW were offered to join the study if they were SARS-CoV-2 naïve, i.e. did not have a previous positive PCR, or detectable anti-RBD IgG before the first vaccine dose. Any HCW who was infected by SARS-CoV-2 during the study, as determined either by positive PCR or by anti-N IgG, was removed from the cohort to a parallel cohort of COVID-19 recovered HCW. All participants were requested to perform a serology test once every 4 weeks and reminders were sent by mails and text messages.

Data on age and sex were available for all 3972 study participants. Overall, 2723 (68%) responded to an online questionnaire regarding comorbidities including immunosuppression and BMI.

In this study, we compared the dynamics of the immune response after the third dose (administered at least five months after the second dose) to that of the second dose. We included 3972 HCW who received a third dose, and had at least one serological test 7 to 140 days after the third dose of vaccine, the number of participants who had tests during the different time points of the study are presented in Figure 1A. The results after the third dose were compared to a cohort of 4868 of the same HCW that had serology testing after the second dose previously described[18]. Serology samples were collected between January 21, 2021 and December 29, 2021.

**Fig 1.**
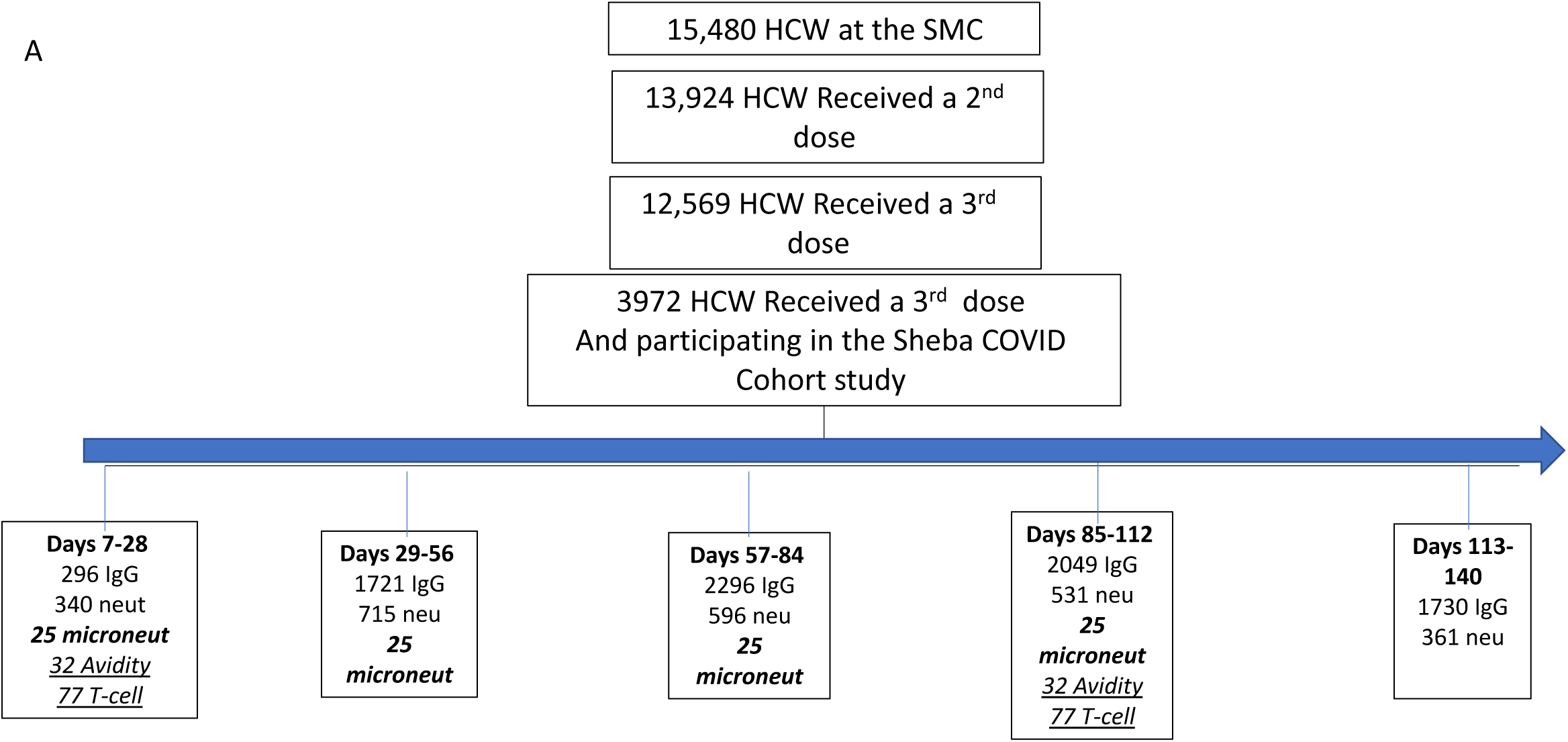
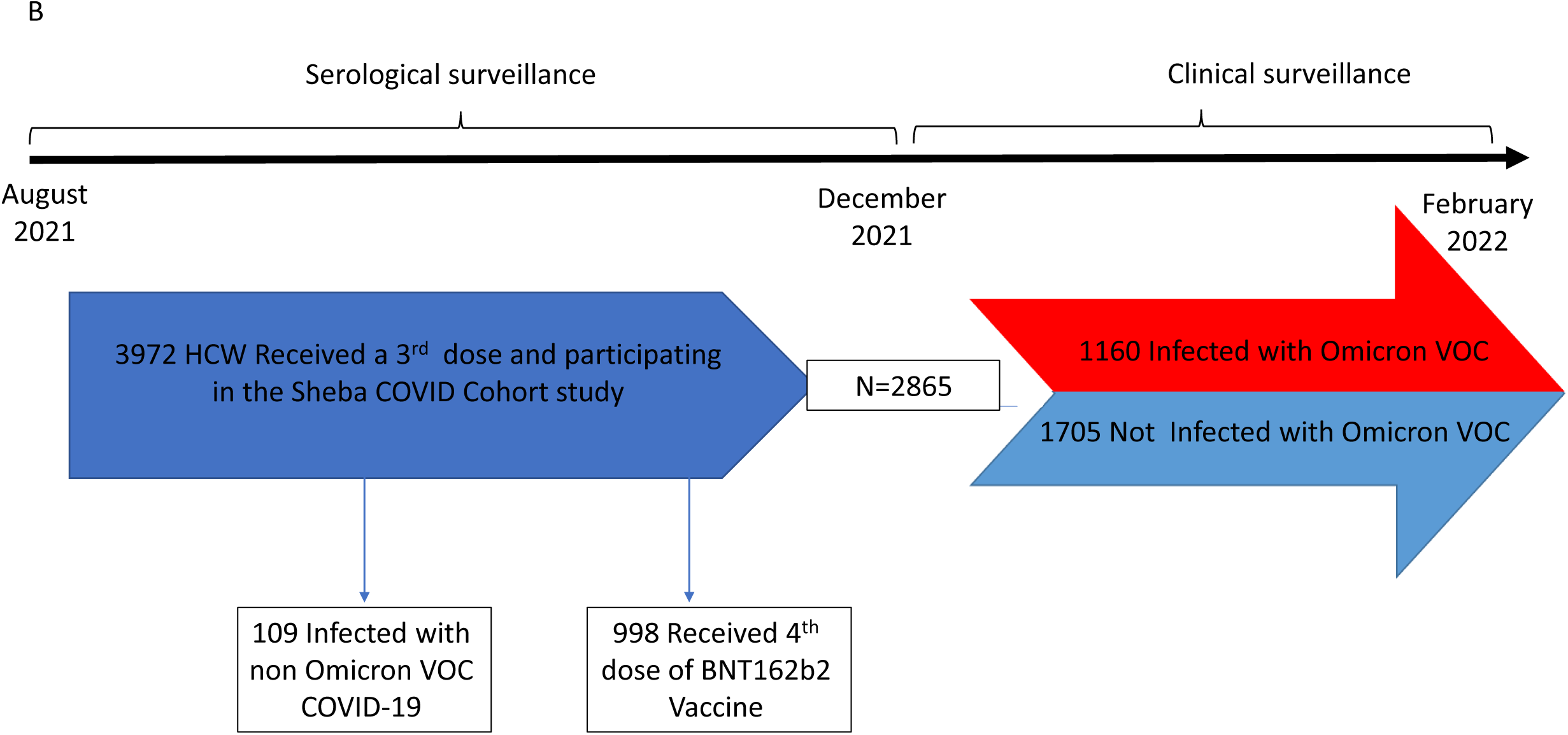
Recruitment of participants, testing and follow-up-. This study involved a prospective cohort of health care workers who had received the third dose of the BNT162b2 vaccine and underwent at least one serologic assay after receipt of the third dose of vaccine. Panel A shows the schedule for serological follow-up. Panel B shows the serological follow-up of those eventually infected with Omicron VOC. Patients who received a fourth dose of vaccine as well as those who had an early breakthrough infection were censored, Serological follow up of those infected obtained after infection was censored as well. IgG-IgG antibodies, neut-neutralizing antibodies, microneut-micro neutralization

To further determine if the rate of waning is a predictor of later breakthrough infections, we followed the clinical outcome of the participants during the Omicron surge, between December 15, 2021 and February 27, 2022. We compared the dynamics of antibody waning after the third dose, between those infected with COVID-19 during the Omicron surge to those who remained uninfected during the follow up period, the study design is depicted in Figure 1B.

### Serological assays

#### IgG antibody assay

Samples from vaccinated participants were tested before receipt of the third dose using the SARS-CoV-2 Receptor Binding Domain (RBD) IgG assay (Beckman-Coulter, CA, U.S.A.), or after receipt of the third dose using the SARS-CoV-2 IgG II Quant (Abbott, IL, USA) test. These commercial tests were performed according to manufacturer’s instructions. To present all IgG Antibody levels in Binding Antibody Units (BAU) per the World Health Organization (WHO) standard measurements we imputed the Abott-based BAU values from the Beckman-Coulter assay results, based on an independent sample of individuals with both Abbott BAU and Beckman-Coulter levels (see Supplementary Methods).

#### Avidity

to measure the quality of IgG antibodies we used urea as a chaotropic reagent and test the strength of interaction between the IgG and the viral antigen (the RBD).

#### SARS-CoV-2 Pseudovirus (psSARS-2) Neutralization Assay

to test the overall neutralizing ability of each serum against the WT virus and specifically to compare with neutralizing levels of the Sheba HCW following two and three vaccine doses we used Pseudovirus (psSARS-2) Neutralization as previously described[22].

#### SARS-CoV-2 micro-neutralization assay

to compare the neutralizing capacity of omicron and delta variants following the third vaccine dose a SARS-CoV-2 micro-neutralization assay with live virus was performed as previously described [17]

#### Memory T cell response

To investigate the memory response we isolated peripheral blood mononuclear cell (PBMC) using Ficoll density gradient centrifugation and using Interferon (IFN)-γ ELISPOT analysis we measured SARS-CoV-2 specific T cell activation as described previously, and detailed in the supplementary[13]

### SARS-CoV-2 detection

During the whole study period, to identify any infection, participants were requested to perform a SARS-CoV-2 test, either RT-PCR (Seegene,Seoul, Korea), or rapid Ag test, in case of any event of exposure to a detected SARS-CoV-2 infected person or in case of development of any potential COVID-19 symptom, including fever, throat pain, headache, myalgia, rhinorrhea, cough or loss of smell or taste. During the Omicron surge (Dec 15, 2021 – end of study: February 28 2022) additionally, all HCW were requested to be tested routinely once a week. All PCR SARS-CoV-2 tests conducted in the hospital or in other settings were reported through a central reporting system.

### Statistical Methods

The analysis of waning of IgG and neutralizing antibody levels was conducted on the same length of follow-up (140 days) for both the second and third vaccine doses. Abbott IgG levels following the second dose were imputed from Beckman-Coulter IgG levels using data on 215 selected serum samples, not included in the HCW cohort, using a cubic polynomial equation in log Beckman-Coulter level (R-squared = 0.92). See Supplementary Materials, Section S6.

Rates of decline in antibody level were analyzed using a linear mixed model, where log antibody level was the dependent variable and each individual’s peak level was modeled as a random effect. Separate models were run for the second and third vaccine doses. The log antibody levels were modeled as constant up to 30 days post-vaccine (peak level), followed by a fixed effect linear decline from 30 days onwards. For neutralizing antibody levels, the decline from 70 days following the second vaccine dose, but not the third dose was found to be slower than the initial rate of decline, and was modeled accordingly; the rates of decline reported here are the initial rates, and all rates are expressed on the original scale as percent reductions per day. Age (<45y, 45-64y, ≥65y) and gender were included as fixed-effect adjusting covariates, and their interactions with peak level and rate of decline were included. For comparisons between doses, estimated average peak levels, rates of decline and the level at day 140 following the third vaccine dose were standardized to the distribution of age and sex of those in the second dose cohort. Standard errors of estimates were model-based for neutralizing antibodies, but for IgG a bootstrap procedure was used to account for the extra uncertainty from imputing Abbott IgG levels for the second dose cohort. See Supplementary materials for further details. To compare the kinetics of infected versus uninfected persons, an extra covariate indicating infection with omicron (yes/no) was entered into the linear mixed model for those who received a third vaccine dose. Interactions between this covariate and peak level, rate of waning, and age group (<65y, ≥65y) were also included. From these models, ratios of average peak levels and rates of waning between infected and uninfected were computed separately for each age group.

### Graphical presentation

Scatter plots of IgG and neutralizing antibody levels since the receipt of the second and third doses were created with the use of GraphPad Prism software, version 9.0 (GraphPad Software). Correlations between IgG and neutralizing antibody levels for each period were assessed by Spearman’s rank correlation. Paired pre-and post-third vaccine dose avidity, neutralization and T cell activation were compared using the Wilcoxon signed-rank test. Statistical analysis was performed using SAS software, version 9.4 (SAS Institute),

## Results

### Study population and serologic assays

In total 8092 samples from 3972 HCW were collected from August 5, 2021 until December 29, 2021. Of these, clinical follow up data were available for 2865 HCW, who were followed during the Omicron surge, between December 15, 2021 and February 27, 2022. (Fig. 1). The demographic characteristics and data on coexisting conditions in the study participants are provided in table S1.

### Comparing the waning of SARS-COV-2 humoral response after the third dose to that after the second dose

Following transformation to BAU (See Supplementary Methods) waning of IgG antibodies against SARS-COV-2 after receipt of the first two doses of BNT162b2 vaccine [9] was compared to the durability after three doses. IgG waning was slower after the third dose of vaccine, 1.32% (95% CI 1.29-1.36) per day vs 2.26% (95% CI 2.13-2.38) per day after the second dose (Figure 2A) (Table 1).

**Table 1.** Geometric mean of IgG and neutralizing antibodies after the second and third doses of vaccine

|  | Second dose<br>(95% CI) | Third dose<br>(95% CI) | Ratio (third dose to<br>second dose)<br>(95% CI) |
| --- | --- | --- | --- |
| ***Peak*** |  |  |  |
| IgG | 1675<br>(1542-1820) | 2801<br>(2727-2878) | 1.67<br>(1.53-1.82) |
| Neut | 611<br>(557-671) | 4315<br>(4051-4595) | 7.06<br>(6.30-7.90) |
| ***Rate of***<br>***waning*** |  |  |  |
| IgG | 2.26%<br>(2.13-2.38) | 1.32%<br>(1.29-1.36) | 0.59<br>(0.56-0.62) |
| Neut | 3.34%<br>(3.11-3.58%) | 1.32%<br>(1.21-1.43%) | 0.39<br>(0.35-0.44) |
| ***At 140 d*** |  |  |  |
| IgG | 140<br>(125-157) | 650<br>(629-673) | 4.64<br>(4.12-5.22) |
| Neut | 132<br>(121-144) | 1001<br>(895-1119) | 7.57<br>(6.57-8.71) |

**Fig 2.**
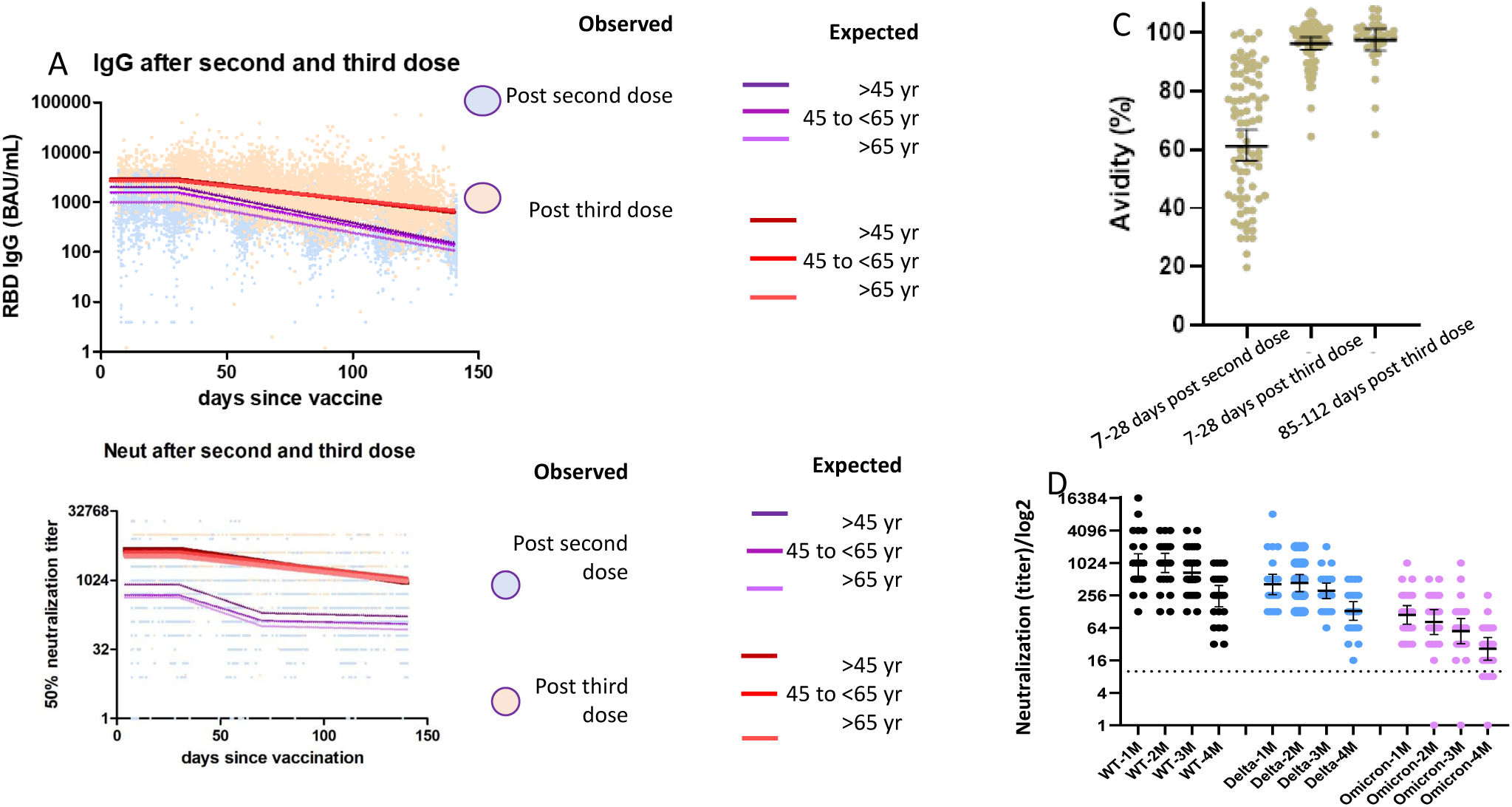
Distribution of antibodies after BNT162b2 vaccines, antibody avidity and micro neutralizaton against VOC-. Panel A shows the distribution of observed IgG antibodies after receiving the second and third dose and the GMT of the expected by a model adjusted by age. Panel B shows the distribution of observed neutralizing antibodies after receiving the second and third dose and the GMT of the expected by a model adjusted by age. Panel C shows the antibody avidity 7-28 days after the second dose of vaccine, 7-28 days after third dose of vaccination and 85-112 das after this dose. Panel D shows the Microneutralization assay against WT, delta and Omicron variants-microneutralization of sera of 25 participants against Wild type (WT), Delta and omicron variants – 1,2,3 and 4 months after third dose of vaccine

Neutralizing antibodies also had a slower rate of decline following the third vaccine dose 1.32% (95% CI 1.21-1.43) vs 3.34% (95% CI 3.11-3.58). Neutralizing antibody kinetics after the third dose were relatively constant and differed from the kinetics after the second dose, where a substantial decline continued beyond 70 days after the second vaccine dose (Figure 2B), (Table 1). Table S7 demonstrates the mixed model analysis of variables associated with IgG and neutralizing antibodies titers and rate of decline.

A strong correlation and constant regression relationship in all study periods was observed between IgG and neutralizing antibodies (Spearman rank correlation between 0.59-0.74) after the third dose (Figure S1).

Avidity was tested on a sub-cohort of 32 participants one and four months after the third vaccine dose and compared to that one month after the second dose. Demographics of this sub cohort are described in table S4. Mean avidity one month after the second dose was 65.7% ±0.2, significantly increasing to 97.4% (SD 0.1) one month after the third vaccine dose and to 98.04% (±0.1) four months after. (Fig 2 C, Table S6)

### Kinetics of SARS-COV-2 cellular immunogenicity after receipt of a third vaccine dose-

T cells activity was tested for 77 participants 7-28 days after the third dose (at peak) and 85-112 days after the third dose (trough). Overall the demographics of this sub cohort were similar to those of the full cohort and its characteristics are described in table S3. Mean T cell activity at peak was 98±5.4 activated T cells/10^6^ PBMC and a decrease to 59±9.3, within 3-5 months. During this time the percentage of T cells non responders increased from 9% to 20% (p=0.0004). IgG and neutralizing antibodies were also tested in this cohort and the dynamics of these were similar to those of the full cohort (Table S6, Figure S2)

### Live microneutralization of Omicron vs. other VOCs

Serum samples from 25 randomly selected individuals with four consecutive monthly samples were tested. The demographics of this sub population are described in table S5. The neutralization GMT titer at peak was 942 (95% CI 585-1518) for WT, 410 (95% CI 266-634) for delta and 111 (95% CI 75-166) for Omicron. For all three tested strains, similar waning within four months were observed (3-4 fold decrease), (Figure 2C, Table S5). Reaching titers of 249 (95% CI158-391), 131 (95% CI 88-197) and 26 (95% CI 16-42) for WT, Delta and Omicron, respectively.

### Omicron breakthrough infections are associated with lower peak IgG levels

To assess if the rate of antibody decline can predict the risk for Omicron breakthrough infections, we followed the clinical outcome of the participants during the Omicron surge (between Dec 15,2021 until Feb 27, 2022). During this period we sequenced 150 random samples taken from HCW and 100% of them where Omicron VOC. A total of 2865 of 3972 HCW, who were not infected before beginning of follow up and did not receive a fourth dose, were followed during this period. See Figure 1B Overall 1160 of 2865 HCW were infected with SARS-COV-2 during the Omicron surge. The duration between third vaccine dose and breakthrough infection varied between subjects and ranged between 82 and 201 days, with a mean of 147.6+-20.6 days (95% CI 106.4-188.8).

Demographics of those infected vs those not infected are described in table S8, those infected were younger than those not infected, with only 4% over 65y compared to 11% among the non-infected, mean age of infected 43.8 (95% CI 43.12-44.45) vs mean of 46.1 (45.46-46.74) among those non infected.. Linear mixed model analysis showed that participants who were infected with COVID-19 at the time of the Omicron VOC surge, had a lower post-3^rd^ dose IgG peak of 2659 (95% CI 2528-2797) vs 3107 (95% CI 2983-3236) (ratio of means between those infected and not infected 0.86, 95% CI 0.80-0.91). (Table 2, Figure 3). Additionally, for participants older than 65y, both the IgG and neutralizing antibodies rate of decline was faster among infected HCW. The rate of decline of IgG antibodies was 1.39% per day (95% CI 1.17-1.62) among those infected vs 0.99% per day (95% CI 0.89-1.10) among those non infected, with a ratio of means of 1.40 95% CI 1.13-1.68. In addition, the rate of decline of neutralizing antibodies was 1.86% per day (95% CI 0.99-2.72) among those infected vs 0.52% per day (95% CI 0.14-0.90) among those not infected, with a ratio of means of 3.58 (95% CI 1.92-6.67).

**Table 2.** Means and rates of decline (% per day) of IgG and neutralizing antibody levels among those eventually infected and those remaining non-infected, and their ratios.

|  | **IgG antibodies**<br>**Ratio of means**<br>**(95% CI\*)** | **Neutralizing**<br>**antibodies**<br>**Ratio of means**<br>**(95% CI)** |
| --- | --- | --- |
| **All ages** |  |  |
| Peak level: infected | 2659 (2528-2797) | 4564 (4047-5149) |
| non-infected | 3107 (2983-3236) | 4654 (4250-5097) |
| ratio | 0.86 (0.80-0.91) | 0.98 (0.84-1.14) |
| Rate of decline (%): infected | 1.33 (1.28-1.38) | 1.40 (1.17-1.62) |
| non-infected | 1.26 (1.22-1.31) | 1.13 (0.97-1.29) |
| ratio | 1.05 (1.00-1.11) | 1.23 (0.97-1.50) |
| **Below 65** |  |  |
| Peak level: infected | 2633 (2503-2769) | 4599 (4061-5209) |
| non-infected | 3118 (2988-3255) | 4784 (4334-5286) |
| ratio | 0.84 (0.79-0.90) | 0.96 (0.82-1.13) |
| Rate of decline (%): infected | 1.32 (1.27-1.38) | 1.33 (1.10-1.55) |
| non-infected | 1.29 (1.24-1.33) | 1.22 (1.05-1.40) |
| ratio | 1.03 (0.97-1.08) | 1.09 (0.85-1.33) |
| **Above 65** |  |  |
| Peak level: infected | 2964 (2300-3818) | 4296 (2853-6469) |
| non-infected | 2981 (2624-3386) | 3853 (3077-4825) |
| ratio | 0.99 (0.75-1.32) | 1.11 (0.70-1.78) |
| Rate of decline (%): infected | 1.39 (1.17-1.62) | 1.86 (0.99-2.72) |
| non-infected | 0.99 (0.89-1.10) | 0.52 (0.14-0.90) |
| ratio | 1.40 (1.13-1.68) | 3.58 (1.92-6.67) |

**Fig 3.**
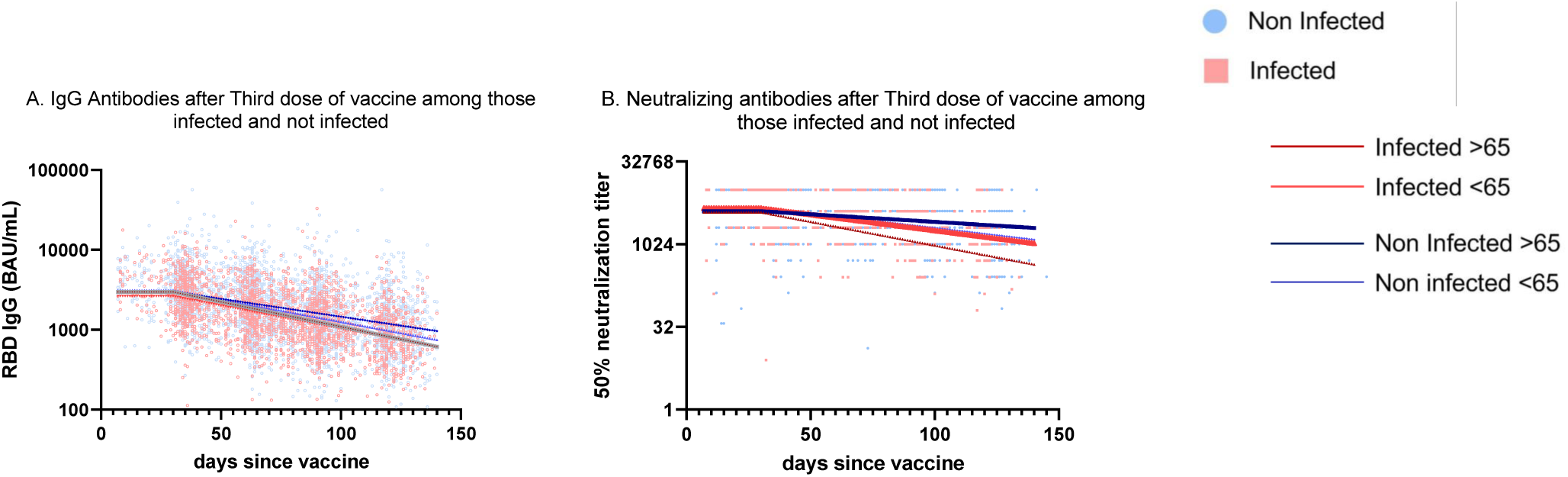
Distribution of antibodies 150 days after receipt of the third doses f BNT162b2 vaccine among those infected and not infected-. Panel A shows the distribution of observed IgG antibodies after receiving the third dose among those eventually infected and those who remained not infected and the GMT of the expected by a model adjusted by age. Panel B shows the distribution of observed neutralizing antibodies after receiving the third dose among those eventually infected and those who remained not infected and the GMT of the expected by a model adjusted by age.

## Discussion

In this prospective longitudinal cohort study, we report significantly slower waning of humoral response following the BNT162b2 third dose vaccine compared to the second dose. We observed that waning of neutralization against Omicron VOC is similar to that against other strains, and is consistently and significantly lower than against Delta and WT during four months of follow-up. Furthermore, we report that Omicron breakthrough infections were associated with lower peak IgG levels and, a faster decay particularly among those over 65y.

Two vaccine doses of BNT162b2 elicit a rapid induction of humoral response[22] followed by significant antibody waning [18,23,24], leading to approximately 30-and 6-fold lower IgG and neutralizing antibodies, respectively, six months after the second dose[18]. The superior humoral response following the third compared to the second dose resulted in an increase in not only the quantity (IgG levels) but, even more importantly, in the quality (avidity) of IgG antibodies [15]. The durability of neutralizing titers following the third vaccine dose may, therefore, be a result of the persistence of high quality IgG antibodies, observed in our study for at least four months. As time passes and possible additional vaccine doses are administrated, future studies should continue to examine the interplay between quantity and quality of IgG antibodies, as these two factors are crucial in determining the humoral response and protection against infection.

We and others have previously demonstrated that neutralizing antibody levels following three vaccine doses are approximately 4 to 10-fold lower against the Omicron compared to WT and the Delta VOC. Here, we report similar waning of direct micro-neutralization against different VOC within 4 months after the third dose. Thus, within a few months of waning, the neutralization efficacy of Omicron may be insufficient to prevent infections or disease. During the Omicron surge in Israel, which began four months after vaccine rollout for the third dose, we witnessed an outstanding number of breakthrough infections even among the vaccinated population [25], raising the suspicion that the immunogenicity toward this variant was substantially decreased despite relatively high IgG and neutralizing antibody levels. Overall, these results suggest that the waning of the booster dose was not the major cause for the high breakthrough infection rate, but rather the relative resistance of Omicron VOC to the humoral response induced even by a full vaccine series (including a booster).

Moreover, we have demonstrated that, at least in individuals over 65y, the rate of immune response waning was associated with Omicron breakthrough infections. Those aged over 65 who eventually had breakthrough infections, had a steeper decline in neutralizing and IgG antibody titers. The reasons for variation in the rate of decline among individuals (of the same age group) has yet to be studied. Nevertheless, a future model that would predict the risk of an individual to be infected, based on peak antibody levels and the rate of their decline, could potentially determine when and who should receive another booster dose.

We have recently shown [26] that a second booster (fourth dose) restored IgG and neutralizing antibody levels to those induced by the first booster[26]. However, we demonstrated that it did not effectively prevent Omicron mild and asymptomatic infections among HCW[26]. Yet, a second booster was efficacious in protecting against severe disease and death compared to one booster dose[27]. Despite the relatively slow waning of the immune response observed here, the lack of protection from infection to emerging SARS-CoV-2 variants suggest that repeated boosters of current available vaccines have reached a limit of protective effect in young and healthy populations, and timing of second and third boosters in such populations should be further considered and studied.

This study has several limitations, first as all participants are health care workers, they are relatively younger and healthier than the general population, thus potentially limiting its generalizability. Second, the study period following waning was relatively short. Further studies to assess waning after longer periods are needed. Furthermore, as participants were not blinded to their serology testing, those who had lower serological markers, could have potentially had a lower threshold for PCR testing and this might have created a bias. Yet, during the study period, due to the outstanding surge in cases, all HCW, regardless of their serological tests, were encouraged to obtain a weekly RT-PCR SARS-COV test in addition to testing after exposure or due to symptoms.

Numerous studies showed that mRNA vaccines played a major role in protecting the world population against COVID-19. We demonstrate here that the humoral response following a third dose is sustained for months with only minor reduction in antibody levels and that infection with omicron VOC is correlated with antibody level and thus, potentially, might be predicted. Nevertheless, our results clearly show that the humoral response generated by vaccination may not be enough to protect against Omicron infection. BNT162b2 booster doses have been proven to be protective against severe disease and mortality, yet if reducing transmissibility and achieving herd immunity is what we are striving for, a different vaccination strategy may be required.

## Data Availability

All data produced in the present study are available upon reasonable request to the authors

