## Supplementary for "Durability of the immune response to a third BNT162b2 dose; five months follow-up"

**Supplementary Materials**

**Table of Contents**

Supplementary Methods S1- PCR testing…………………………………………………….2

Supplementary Methods S2- Inclusion criteria for selecting the neutralizing antibody group…………………………………………………………………………………………..3

Supplementary Methods S3- antibody detection testing …………………………………….4-5

Supplementary Methods S4- SARS-COV-2 microneutralization…………………………..………..……….6

Supplementary Methods S5- Memory immune response……………………………..……….7

Supplementary Method S6 – Imputation of Binding Antibody Units and IgG Linear Mixed Model ………………………………………………...…….……………………….…………8

Supplementary Methods S7- Neutralizing antibody linear mixed model……………..…....9-10

Supplementary Results S1 – IgG Linear Mixed Model………………….…………………….11

Supplementary Results S2 – Neutralizing antibody Linear Mixed Model………………...….12

Supplementary Table S1- Baseline characteristics of the IgG study population……………..….....13

Supplementary Table S2- Baseline characteristics of the neutralizing antibodies study population……………………………………………………………………………………………………………………….....14

Supplementary Table S3- Baseline characteristics of the T cell activation population…………..15

Supplementary Table S4 Baseline characteristics of the avidity study population………...……..15

Supplementary Table S5- Baseline characteristics of the microneutralization study population……………………………………………………………………………………………………………………..….16

Supplementary Table S6- observed results of study population by the different time periods since third dose of vaccination……………………………………………………………………………………………16

Supplementary Table S7- Mixed Model Analysis of Variables Associated with IgG and Neutralizing Antibody Titers after receipt of the Second Vaccine Dose……………………………………………………………17

Supplementary Table S8- Baseline characteristics of those eventual infected Vs those non infected………………………………………………………….…………………………………………………………………..18

Supplementary Table S9- Computer-based questionnaire………….………………...……..19

Supplementary Table S10- Variable definitions………….…………………..…............…….20

Supplementary Figure S1- Correlation between Neutralizing antibodies titers and IgG antibodies titers by month-…………………………………………………………………....21

Supplementary Figure S2: Immunogenicity of a sub cohort of 77 participants- ……………..22

Supplementary References…………………………………...……………………………….23

**Supplementary Methods S1 - PCR testing**

Hospital personnel were tested in several scenarios: upon every symptom suspected to be COVID-19, following exposure to a positive COVID-19 contact (hospital or community contacts).

For quantitative RealTime-PCR (qRT-PCR), nasopharyngeal swabs were placed in 3mL of universal transport medium (UTM) or viral transport medium (VTM). Test was performed according to manufacturers' instructions on various platforms: Allplex™ 2019-nCoV (Seegene, S. Korea), NeuMoDx™ SARS-CoV-2 assay (NeuMoDx™ Molecular, Ann Arbor, Michigan), Xpert®, Xpress SARS-CoV-2 (Cepheid, Sunnyvale, CA, USA).

**Supplementary Method S2- Inclusion criteria for selecting the neutralizing antibody subgroup**

1. Age ≥65
2. Body mass index ≥30
3. Pregnancy
4. Allergy
5. Hypertension
6. Diabetes
7. Dyslipidemia
8. Heart disease
9. Lung disease
10. Kidney disease
11. Liver disease
12. Autoimmune disease
13. Immunosuppression

Additionally, 50% of healthy health care workers were randomly selected for the neutralizing antibody subgroup.

**Supplementary Methods S3- antibody detection**

**SARS-CoV-2 IgG Assay**

Samples from vaccinated participants were tested before receipt of the third dose using the SARS-CoV-2 Receptor Binding Domain (RBD) IgG assay (Beckman-Coulter, CA, U.S.A.), or after receipt of the third dose using the SARS-CoV-2 IgG II Quant (Abbott, IL, USA) test. These commercial tests were performed according to manufacturer's instructions. To present all IgG Antibody levels in Binding Antibody Units (BAU) per the World Health Organization (WHO) standard measurements we imputed the Abott-based BAU values from the Beckman-Coulter assay results, based on an independent sample of individuals with both Abbott BAU and Beckman-Coulter levels .

**Avidity –** to measure the quality of IgG antibodies we used urea as a chaotropic reagent and test the strength of interaction between the IgG and the viral antigen (the RBD). Specifically, a 96 well microtiter Polysorb plate (Nunc, Thermo, Denmark) was coated overnight at 4°C with 50μl per well of 1μg/ml of RBD antigen. After blocking with 5% skimmed milk at 25°C for 60 minutes, serum samples were be diluted 1:100, 1:400 and 1:1000 with 3% skimmed milk and added to antigen coated wells. The plate was incubated at 25°C for 120 minutes, and following washing each sample,incubated either with the addition of 6M urea or PBS for 10 min. After washing, a goat anti-human IgG horseradish peroxidase (HRP) conjugate (Jackson ImmunoResearch, PA, USA Code: 109-035-088) (diluted 1:15000) was be added to each well for 60 min. After washing, incubation of TMB Substrate Solution (Abcam) for 5 min and the addition of stop solution (2N HCl), the OD of each well was measured at 450nm using a micro-plate reader (Sunrise, Tecan). Avidity index was calculated as the ratio (in percentage) between sample OD with 6M urea and sample OD with PBS.

**SARS-CoV-2 Pseudovirus (psSARS-2) Neutralization Assay**

to test the overall neutralizing ability of each serum against the WT virus and specifically to compare with neutralizing levels of the SHEBA HCW following one, two and three vaccine doses we used Pseudovirus (psSARS-2) Neutralization as previously described (references). SARS-CoV-2 Pseudo-virus (psSARS-2) Neutralization Assay was performed using a propagation-competent VSV-spike which was shown to be highly correlative to authentic SARS-CoV-2 virus micro-neutralization assay. Following titration, 100 focus forming units (ffu) of psSARS-2 was incubated with 2-fold serial dilution of heat inactivated (56°C for 30 min) tested sera. After incubation for 60 min at 37°C, virus/serum mixture was transferred to Vero E6 cells that have been grown to confluency in 96-well plates and incubated for 90 min at 37°C. After the addition of 1% methyl cellulose in dulbecco's modified eagle's medium (DMEM) with 2% of fetal bovine serum (FBS), plates will be incubated for 24hr and 50% plaque reduction titer was calculated by counting green fluorescent foci using a fluorescence microscope. Sera not capable of reducing viral replication by 50% at 1 to 16 dilution or below was considered non- neutralizing.

**Supplementary Methods S4- SARS-CoV-2 micro-neutralization**

**-**to compare the neutralizing capacity of omicron and delta variants following the third vaccine dose a SARS-CoV-2 micro-neutralization assay with live virus was performed as previously described ^1,2^.VERO-E6 cells at concentration of 20*103/well were seeded in sterile 96-wells plates with 10% FCS MEM-EAGLE medium, and stored at 37⁰C for 24 hours. One hundred TCID50 of Wild Type, Beta, Delta and Omicron SARS-CoV-2 isolates were incubated with inactivated sera diluted 1:10 to 1:16,384 in 96 well plates for 60 minutes at 33ºC. Virus-serum mixtures were added to the Vero E-6 cells and incubated for five days at 33ºC after which Gentian violet staining (1%) was used to stain and fix the cell culture layer. Neutralizing dilution of each serum sample was determined by identifying the well with the highest serum dilution without observable cytopathic effect. A dilution equal to 1:10 or above was considered neutralizing.

**Supplementary Methods S5- Memory immune response -**

To investigate the memory response we isolated peripheral blood mononuclear cell (PBMC) using Ficoll density gradient centrifugation and analyzed T cell activation as desctibed previously ^3^.

T cell activation was be assessed by IFN-γ ELISpot assay. Specifically, IFN- γ -secreting cells were enumerated using Elispot IFN-γ kits (IFN-γ kit, AID Autoimmun Diagnostika GmbH, Strassberg, Germany) according to manufacturer instructions. For antigen stimulation, 50 μl of SARS-CoV-2 peptide pools (S-complete, Miltenyi Biotech) was used. Test medium was used as negative control and Phytohaemagglutinin (PHA) was used as positive control. IFN-γ-secreting cells frequency were quantified using the AID ELISpot Reader (Strassberg, Germany). The unspecific background (mean SFU from negative control wells) was subtracted from experimental readings.

**Supplementary Method S6 – Imputation of Binding Antibody Units and IgG Linear Mixed Model**

**S6.1 Imputation of Binding Antibody Units based on Beckman-Coulter Assay Results**

We developed a method for imputing Abbott IgG levels from Beckman-Coulter IgG levels using data on 215 selected serum samples, taken from individuals who had not received a booster dose and were not included in the HCW cohort, and were measured by both methods. We fitted a cubic polynomial regression model where log (to the base e) of IgG measured in BAU units by the Abbott kit was regressed on the log (to the base e) of IgG measured by the Beckman-Coulter kit, its squared value, and its cubed value. The fitted regression equation, using the glm procedure in R, was:

logIGG_Abbott =

4.506 + 0.6634 × logIGG_Beckman - 0.0852× (logIGG_Beckman)^2^ + 0.0403× (logIGG_Beckman)^3^

**Output from the glm procedure in R**

```
Coefficients:
              Estimate Std. Error t value Pr(>|t|)
(Intercept)   4.506257   0.039815 113.180  < 2e-16 ***
logBeck       0.663432   0.042540  15.595  < 2e-16 ***
I(logBeck^2) -0.085175   0.020513  -4.152 4.78e-05 ***
I(logBeck^3)  0.040341   0.007083   5.695 4.10e-08 ***
---
Signif. codes:  0 ‘***’ 0.001 ‘**’ 0.01 ‘*’ 0.05 ‘.’ 0.1 ‘ ’ 1

Residual standard error: 0.4377 on 211 degrees of freedom
Multiple R-squared:  0.9187,	Adjusted R-squared:  0.9176
```

­­­­­­­­­­­­­­­­­­­­­­­­­­­­A simple linear model has an R-squared of 0.905, compared to 0.919 for the cubic polynomial. One can see from the figure below that the cubic polynomial fits the data much better at the lower and upper ends of the scale.

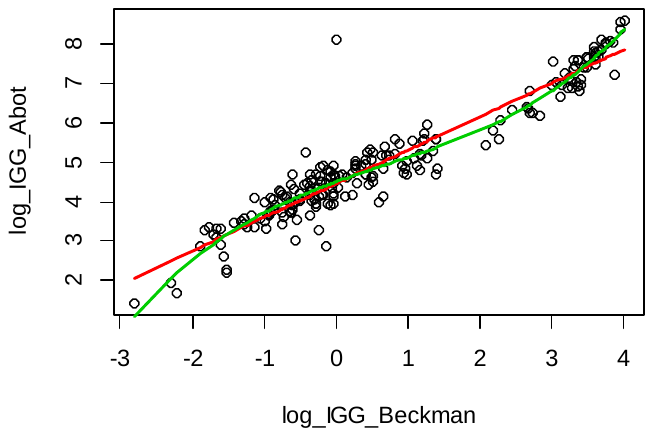

The regression equation shown above was used to impute the values of Abbott BAU for samples taken after the second vaccine dose. In order to avoid extrapolation, any Beckman-Coulter IgG value that was lower than the minimum value of the calibration sample (0.06) was censored to that minimum (occurring in 0.1% of the sample) and any value above the maximum value (55.29) was censored to that maximum (occurring in 2.2% of the sample). Abbott BAU levels following the third vaccine dose were measured directly and did not require imputation.

**S6.2 Linear Mixed Model for IgG Level Kinetics Following Second and Third Vaccine Doses**

We modeled the natural log-transformed IgG in a mixed effects linear model with subject level random effects. Separate models were run for the second and third vaccine doses. We fit a linear slope from 30 days after vaccination onwards. IgG tests taken before 30 days were used to estimate the “peak” IgG attained. We fitted a model that included age group (<45, 45-64, ≥65), sex, time (measured in days from day 30 post-vaccine onwards; all measurements before day 30 were counted as day 0), and all age-sex-time interactions (including the 3-way interaction) as fixed effects, and subject-level random intercept. For the third dose, the three-way interaction between age, sex and time was small and not statistically significant, and therefore omitted.

Peak IgG levels, rates of decline and level at day 140 following vaccine for each participant’s age-sex profile were estimated from the coefficients obtained from the model fits and averaged over the cohorts. For comparisons between the kinetics following the second and third doses, the averages for the third vaccine dose were standardized to the distribution of age and sex of those in the second dose cohort. Standard errors of estimates were based on a bootstrap procedure to account for the extra uncertainty from imputing Abbott IgG levels for the second dose cohort. In this procedure, each bootstrap run included a bootstrap sample from the imputation dataset that was used to re-calculate the coefficients of the cubic equation; the new equation was then applied to a bootstrap sample of the second cohort to impute the BAU values and the linear mixed model was run on these values.

**S6.3 Comparison between Omicron-Infected and Non-Infected persons with regard to IgG Level Kinetics Following Third Vaccine Dose**

To compare the kinetics of omicron-infected versus uninfected persons, an extra covariate indicating infection with omicron (yes/no) was entered into the linear mixed model for those who received a third vaccine dose, and interactions between age (<65, ≥65), sex, time and omicron infection were tested and retained if significant at the 5% level. When a higher-order interaction was retained all its lower-order interactions were also retained. In this way, we included in the model all main effects and the following interactions: age-omicron-time; age-time; omicron-time; sex-time. From the parameter estimates of this model, ratios of average peak levels and rates of waning between omicron-infected and uninfected persons were computed separately for each age group. Standard errors of the ratios were computed using the delta method.

**Supplementary Method S7 – Neutralizing antibody Linear Mixed Model**

We modeled log-transformed (to the base 2) neutralizing antibody (NeutAb) using a linear mixed effects model similar to the model used for the IgG analysis (Supplementary Methods S6.2). Separate models were run for the second and third vaccine doses. The model included subject-level random effects. For the second dose, NeutAb kinetics over time were different from IgG kinetics. The decline started from 30 days after vaccination as in the IgG data (see Supplementary Methods S6.2). However, the decline lasted only up to ~70 days after vaccination, and then stabilized to a much slower decline. The change point at 70 days was chosen based on inspection of the fit of previous data without the change point, and examining AIC for models with a change point at 60, 70, 80 and 90 days. Therefore, we included in the model a separate slope from day 70 to day 140 post-vaccine. For the third dose, the slope after day 70 was not more gradual than up to day 70 and the decline was therefore modeled as a linear slope from day 30 onwards (as for IgG levels). Other main effects and interactions, including interaction between fixed-effect factors with time in the period from day 30 to day 70, were the same as in the model for IgG levels, except that for the third dose, the interaction between age and sex was small and non-significant, and therefore omitted.

Average peak NeutAb levels, rates of decline and level at day 140 following vaccine were estimated and compared between doses using the same methods as described in Supplementary Methods S6.2. Standard errors of estimates were model-based, and those of ratios were based on the delta method.

To compare the kinetics of omicron-infected versus uninfected persons, the same methods were used as for IgG levels (see Supplementary Methods S6.3), and the same interaction terms as for the IgG model were retained. From the parameter estimates of this model, ratios of average peak levels and rates of waning between omicron-infected and uninfected persons were computed separately for each age group. Standard errors of these ratios were computed using bootstrap methods, because, with the smaller numbers in this analysis, the asymptotic delta method was thought less reliable.

**Supplementary Results S1 – IgG Linear Mixed Models**

The results of fitting the linear mixed model for the IgG level kinetics following the second and third doses are shown below.

1. Model fixed parameter estimates for natural log IgG levels following **second** dose

(se denotes the standard error obtained by the bootstrap sampling; pval denotes the p-value obtained from the z-test based on est/se)

```
Fixed effects: ligg ~ agec * male * days30

                          est     se   pval
(Intercept)            7.6537 0.0491 0.0000
agec45-64             -0.2655 0.0346 0.0000
agec65+               -0.6006 0.1018 0.0000
male                  -0.1099 0.0470 0.0193
days30*               -0.0239 0.0008 0.0000
agec45-64:male        -0.0802 0.0695 0.2484
agec65+:male          -0.2567 0.1654 0.1207
agec45-64:days30       0.0017 0.0003 0.0000
agec65+:days30         0.0041 0.0009 0.0000
male:days30            0.0016 0.0004 0.0002
agec45-64:male:days30 -0.0011 0.0006 0.0533
agec65+:male:days30   -0.0026 0.0013 0.0448

* days 30 is defined as taking value 0 until day 30 and increasing from 0 to 110
linearly until 140.
```

2. Model fixed parameter estimates for natural log IgG levels following **third** dose

(Standard errors obtained from the model; p-values based on the t-test)

```
Fixed effects: ligg ~ agec * male + days30 * agec + male * days30

                     Value  Std.Error   DF  t-value p-value
(Intercept)       7.964272 0.02449155 4105 325.1845  0.0000
agec45-64        -0.029576 0.03284721 3919  -0.9004  0.3680
agec65+          -0.002495 0.05152954 3919  -0.0484  0.9614
male             -0.005061 0.04868796 3919  -0.1040  0.9172
days30           -0.014259 0.00025796 4105 -55.2775  0.0000
agec45-64:male   -0.044696 0.06771464 3919  -0.6601  0.5093
agec65+:male     -0.180259 0.08295398 3919  -2.1730  0.0298
agec45-64:days30  0.000880 0.00030912 4105   2.8453  0.0045
agec65+:days30    0.001064 0.00039535 4105   2.6922  0.0071
male:days30       0.001816 0.00032140 4105   5.6515  0.0000
```

3. Model fixed parameter estimates for comparing kinetics between **omicron-infected and uninfected persons** following the **third** dose. (Standard errors obtained from the model; p-values based on the t-test)

```
                    Value Std.Error   DF  t-value p-value
(Intercept)        8.0524    0.0232 2860 346.6659  0.0000
age65             -0.0394    0.0689 2860  -0.5717  0.5676
male              -0.0372    0.0405 2860  -0.9187  0.3583
omic              -0.1692    0.0338 2860  -5.0105  0.0000
days30            -0.0134    0.0002 2645 -56.4218  0.0000
age65:omic         0.1634    0.1486 2860   1.0998  0.2715
male:days30        0.0021    0.0004 2645   4.7262  0.0000
omic:days30       -0.0004    0.0004 2645  -0.9844  0.3250
age65:days30       0.0027    0.0006 2645   4.3296  0.0000
age65:omic:days30 -0.0037    0.0014 2645  -2.7325  0.0063
```

**Supplementary Results S2 – Neutralizing Antibody Linear Mixed Models**

The results of fitting the linear mixed model for the neutralizing antibody (NeutAb) level kinetics following the second and third doses are shown below.

1. Model fixed parameter estimates for log (base 2) NeutAb levels following **second** dose

(Standard errors obtained from the model; p-values based on the t-test)

```
Fixed effects: lneut ~ agec * male * days30a + days70

                        Value Std.Error   DF  t-value p-value
(Intercept)            9.9773    0.1143 2472  87.2604  0.0000
agec45-64             -0.9978    0.1600 1222  -6.2364  0.0000
agec65+               -0.9442    0.1764 1222  -5.3533  0.0000
male                  -0.9563    0.2896 1222  -3.3021  0.0010
days30a*              -0.0559    0.0027 2472 -20.9726  0.0000
days70                -0.0035    0.0010 2472  -3.4490  0.0006
agec45-64:male         0.5844    0.4057 1222   1.4405  0.1500
agec65+:male           0.2030    0.3621 1222   0.5606  0.5752
agec45-64:days30       0.0120    0.0035 2472   3.4765  0.0005
agec65+:days30         0.0056    0.0040 2472   1.3977  0.1623
male:days30            0.0165    0.0062 2472   2.6670  0.0077
agec45-64:male:days30 -0.0246    0.0089 2472  -2.7716  0.0056
agec65+:male:days30   -0.0201    0.0080 2472  -2.5076  0.0122

* days30a is defined as taking value 0 until day 30, increasing from 0 to 40
linearly until day 70, and then remaining constant at 40 from day 70 to 140.
```

2. Model fixed parameter estimates for log (base 2) IgG levels following **third** dose

(Standard errors obtained from the model; p-values based on the t-test)

```
Fixed effects: ligg ~ days30 * agec + male * days30

                   Value Std.Error   DF  t-value p-value
(Intercept)      12.3447    0.0832 1602 148.2980  0.0000
agec45-64        -0.2311    0.1016  974  -2.2754  0.0231
agec65+          -0.4501    0.1156  974  -3.8941  0.0001
days30*          -0.0231    0.0015 1602 -15.4331  0.0000
male             -0.4456    0.0951  974  -4.6857  0.0000
agec45-64:days30  0.0045    0.0018 1602   2.5022  0.0124
agec65+:days30    0.0045    0.0020 1602   2.2191  0.0266
days30:male       0.0058    0.0016 1602   3.5713  0.0004

* days 30 is defined as taking value 0 until day 30 and increasing from 0 to 110
linearly until 140.
```

3. Model fixed parameter estimates for comparing kinetics between **omicron-infected and uninfected persons** following the **third** dose. (Standard errors obtained from the model; p-values based on the t-test)

```
                    Value Std.Error  DF  t-value p-value
(Intercept)       12.3019    0.0773 936 159.2258  0.0000
male              -0.3782    0.1277 591  -2.9615  0.0032
days30            -0.0193    0.0014 936 -13.8837  0.0000
age65             -0.2308    0.1819 591  -1.2692  0.2049
omic              -0.0569    0.1170 591  -0.4865  0.6268
male:days30        0.0075    0.0024 936   3.1442  0.0017
days30:age65       0.0086    0.0031 936   2.7256  0.0065
age65:omic         0.2137    0.3636 591   0.5879  0.5568
days30:omic       -0.0015    0.0021 936  -0.7261  0.4680
days30:age65:omic -0.0180    0.0074 936  -2.4289  0.0153
```

**Supplementary Table S1 – Baseline characteristics of the IgG study population**

|  | Second dose IgG cohort (n=4868) | Third dose IgG cohort (n=3972) |
| --- | --- | --- |
| Number of tests |  |  |
| 1 | 2237 (46%) | 1795 (45%) |
| 2 | 553 (11%) | 802 (20%) |
| 3 | 297 (6%) | 841 (21%) |
| 4 | 285 (6%) | 499 (13%) |
| 5 | 1496 (31%) | 35 (1%) |
| Gender- male (%) | 1310 (27%) | 996 (25.1%) |
| Age-mean+-SD | 46.9+-13.7 | 48.5+-14.1 |
| 18-45 | 2241 (46%) | 1704 (43%) |
| 45-65 | 2072(43%) | 1727 (43%) |
| Over 65 | 555 (11%) | 541 (14%) |
| BMI | 25.5 (+-4.6) | 21.3+-10.6 |
| <25 | 1948 (52%) | 1582 (58%) |
| 25-30 | 1216 (32%) | 750 (28%) |
| >30 | 446 (12%) | 390 (14%) |
| Immunosuppressed | 41 (1%) | 25 (1%) |
| Number of comorobidities |  |  |
| 0 | 3001 (79%) | 2029 (75%) |
| 1 | 568 (15%) | 467 (17%) |
| >2 | 239 (6%) | 227 (8%) |

**Supplementary Table S2 – Baseline characteristics of the neutralizing antibodies study population**

|  | Second dose neutralizing cohort (n=1269) | Third dose neutralizing cohort (n=874) |
| --- | --- | --- |
| Number of tests |  |  |
| 1 | 314 (25%) | 311 (36%) |
| 2 | 179 (14%) | 195 (22%) |
| 3 | 182 (14%) | 186 (21%) |
| 4 | 153 (12%) | 150 (17%) |
| 5 | 306 (24%) | 32 (4%) |
| Gender- male (%) | 310 (24%) | 253 (29%) |
| Age-mean | 52.7+-14.2 | 52.5+- 14.0 |
| 18-45 | 398 (31%) | 280 (32%) |
| 45-65 | 527 (42%) | 401(34%) |
| Over 65 | 344 (27%) | 193 (34%) |
| BMI | 26.9+-5.2 | 23.1+-9 |
| <25 | 488 (43%) | 413 (54%) |
| 25-30 | 325 (29%) | 229 (30%) |
| >30 | 326 (29%) | 120 (16%) |
| Immunosuppressed | 28 (2%) | 8 (1%) |
| Number of comorobidities |  |  |
| 0 | 785 (62%) | 517 (68%) |
| 1 | 233 (18%) | 151 (20%) |
| >2 | 131 (10%) | 94 (12%) |

**Supplementary Table S3 – Baseline characteristics of the T cell activation study population**

| T Cell cohort n=77 |  |
| --- | --- |
| Male | 22/77 (29%) |
| Age | 52+-13 |
| <45 | 27 (35%) |
| 45-60 | 29 (38%) |
| >60 | 21 (27%) |
| BMI | 25.8+-5 |
| Number of comorbidities |  |
| 0 | 50/75 (67%) |
| 1 | 13/75 (17%) |
| 2+ | 12/75 (16%) |
| Immunosuppression | 2/75 (3%) |

**Supplementary Table S4 – Baseline characteristics of the avidity study population**

| Avidity cohort | n=32 |
| --- | --- |
| Male | 3 (9%) |
| Age | 51.5+-7.6 |
| <45 | 8 (25%) |
| 45-60 | 19 (59%) |
| >60 | 5 (16%) |
| BMI | 25.0+-7.2 |
| Number of comorbidities |  |
| 0 | 20 (63%) |
| 1 | 4 (13%) |
| 2+ | 8 (25%) |
| Immunosuppression | 2 (6%) |

**Supplementary Table S5 – Baseline characteristics of the microneutralization study population**

| Microneutralization | 1 month n=25 | 2 months n=25 | 3 months  N=25 | 4 months  N=25 |
| --- | --- | --- | --- | --- |
| Age | 45.2+-15.2 | 45.3+-14.1 | 45.3+-12 | 45.5+-24.1 |
| Male % | 6 (24%) | 5 (20%) | 4(16%) | 4 (16%) |
| Immunosuppression | 0/18 (0%) | 0/18 (4%) | 1/17 (6%) | 0/17 (0%) |
| BMI | 24.7+-3.4 | 25.4+-3.7 | 27.2+-4.9 | 28.1+-5.3 |
| Number of comorbidities |  |  |  |  |
| 0 | 14 (78%) | 14 (78%) | 13 (76%) | 8 (47%) |
| 1 | 2 (11%) | 4 (22%) | 2 (12%) | 6 (35%) |
| 2+ | 2 (11%) |  | 2 (12%) | 3 (18%) |

**Supplementary Table S6– Observed results of study population by the different time periods since third dose of vaccination**

| Days |  |  | **7-28** | 29-56 | 57-84 | **85-112** | 113-140 |
| --- | --- | --- | --- | --- | --- | --- | --- |
| IgG |  | N | 296 | 1721 | 2296 | 2049 | 1730 |
|  |  | GMT | 3482 | 2526 | 1604 | 1186 | 853 |
|  |  | SEM | 158 | 76 | 44 | 48 | 55 |
| Neut |  | N | 340 | 715 | 596 | 531 | 361 |
|  |  | GMT | 3819 | 3942 | 3062 | 2071 | 1098 |
|  |  | Geometric SD | 205 | 138 | 148 | 133 | 135 |
| *Sub-cohort with t cells* |  |  |  |  |  |  |  |
| IgG |  | N | 71 |  |  | 71 |  |
|  |  | GMT | 2342 |  |  | 978 |  |
|  |  | Geometric SD | 2.3 |  |  | 2.5 |  |
| Neut |  | N | 73 |  |  | 73 |  |
|  |  | GMT | 3722 |  |  | 2134 |  |
|  |  | Geometric SD | 2.53 |  |  | 3.12 |  |
| T cells |  |  |  |  |  |  |  |
|  |  | N | 79 |  |  | 79 |  |
|  |  | Mean | 98 |  |  | 59 |  |
|  |  | SEM | 5.4 |  |  | 9.3 |  |
|  |  | Percent non responders | 7/79 (9%) |  |  | 16/79  (20%) |  |
| Avidity |  | N | 32 |  | 32 |  |  |
|  |  | Mean | 97.41 |  | 98.04 |  |  |
|  |  | SD | 0.103 |  | 0.095 |  |  |
| Neutralization |  | N | 25 | 25 | 25 | 25 |  |
|  | WT (95% CI) |  | 942.3 (585-1518) | 1024 (678-1548) | 675.6 (443-1029) | 248.7 (158-391) |  |
|  | Delta (95% CI) |  | 410.1 (266-634) | 433.5 (299-629) | 310.8 (225-430) | 131.8 (88-197) |  |
|  | Omicron (GMT+-SEM) |  | 111.4 (75-166) | 82.14 (48-140) | 55.72 (32-96) | 26.14 (16-42) |  |

. **Supplementary Table S7** Mixed Model Analysis of Variables Associated with IgG and Neutralizing Antibody Titers after receipt of the Third Vaccine Dose

| Variable | Peak Titer | | Rate of waning | | 140 days post-vaccine Titer | |
| --- | --- | --- | --- | --- | --- | --- |
|  | IgG | Neut | IgG | Neut | IgG | Neut |
| Age group |  |  |  |  |  |  |
| <45 yr | Reference | Reference | Reference | Reference | Reference | Reference |
| 45 to >65 yr | 0.97  (0.91-1.03) | 0.85 (0.74-0.98) | 100.1%  (100.0-100.1%) | 100.3% (100.1-100.6%) | 1.07  (1.00-1.14) | 1.20 (1.04-1.37) |
| >= 65 yr | 1.00  (0.90-1.10) | 0.73 (0.63-0.86) | 100.1%  (100.0-100.2%) | 100.3%  (100.0-100.6%) | 1.12 (1.01-1.24) | 1.03 (0.88-1.21) |
| Sex |  |  |  |  |  |  |
| Female | Reference | Reference | Reference | Reference | Reference | Reference |
| Male | 0.99 (0.90-1.09) | 0.73 (0.65-0.84) | 100.2%  (100.1-100.2%) | 100.4%  (100.2-100.6%) | 1.22  (1.10-1.34) | 1.14 (1.00-1.30) |

**Supplementary Table S8– Baseline characteristics of Those eventually infected Vs those non infected.**

|  | infected | Not Infected |
| --- | --- | --- |
| N | 1160 | 1705 |
| Gender- male (%) | 225 (19%) | 377 (22%) |
| Age (y) |  |  |
| Mean | 43.8 (95% CI 43.12-44.45) | 46.1 (95% CI 45.46-46.74) |
| 18-44 | 587 (51%) | 780 (46%) |
| 45-64 | 526 (45%) | 742 (44%) |
| ≥65 | 47 (4%) | 183 (11%) |
| Neutralizing antibodies measured | 222 (19%) | 374 (22%) |

**Supplementary Table S9- Computer-based questionnaire**

|  | **Question** | **Answer1** | **Answer2** |
| --- | --- | --- | --- |
| **1** | What is your date of birth? |  | |
| **2** | What is your gender? | Male | Female |
| **3** | What is your current height in m? |  | |
| **4** | What is your current weight in kg? |  | |
| **5** | Did you perform an IgG assay before receiving the first dose of the vaccine? | Yes | No |
| **6** | Do you have high blood pressure disease (systolic blood pressure above 140) treated with medication? | Yes | No |
| **7** | Do you have dyslipidemia (total cholesterol above 200 or LDL cholesterol above 160) treated with medication? | Yes | No |
| **9** | Do you have autoimmune disease treated with medication? | Yes | No |
| **10** | Do you have diabetes (HbA1C>6.5 or fasting blood sugar>126) treated with medication? | Yes | No |
| **11** | Do you have heart disease treated with medication? | Yes | No |
| **12** | Do you have lung disease as asthma, COPD, lung fibrosis treated with medication/s? | Yes | No |
| **13** | Do you have any coagulation disorder resulting in hemorrhage or thrombosis treated with medication? | Yes | No |
| **14** | Are you immunosuppressed (organ transplantation, biologic therapy, chemotherapy, steroids, splenectomy, or HIV)? | Yes | No |
| **15** | Have you ever had a serious allergic reaction (anaphylaxis) that required immediate treatment? | Yes | No |
| **16** | Do you have liver disease as cirrhosis, hepatitis, liver cancer, metabolic disorder? | Yes | No |
| **17** | Do you have kidney disease (creatinine>1.2 or GFR<60) treated with medication? | Yes | No |
| **18** | Are you pregnant (confirmed by a beta HCG blood test and ultrasound fetal heartbeats detection)? | Yes | No |

The questionnaire was reviewed and approved by the Institutional review board of the Sheba Medical Center**.**

IgG=Immunoglobulin G; BMI=Body mass index; Kg=kilogram. M=meter; LDL=low-density lipoproteins; HbA1C=hemoglobin A1C; COPD= chronic obstructive pulmonary disease; HIV=human immunodeficiency; GFR=Glomerular filtration rate.

**Supplementary Table S10- Variable Definitions**

| **Variable** | **Values** | **Definitions** | **Timing** |
| --- | --- | --- | --- |
| **Outcomes** | | | |
| IgG at the peak period | Continuous (S/CO) | SARS-CoV-2 Receptor Binding Domain (RBD) Immunoglobulin G (IgG) assay (Beckman-Coulter, CA, U.S.A.) | During the peak period (days 7-28 after the third vaccination) |
| NeutAb at the peak period | Continuous (50% titer) | SARS-CoV-2 Pseudo-virus (psSARS-2) Neutralization Assay | During the peak period (days 7-28 after the third vaccination) |
| IgG in the EoS | Continuous (S/CO) | SARS-CoV-2 Receptor Binding Domain (RBD) Immunoglobulin G (IgG) assay (Beckman-Coulter, CA, U.S.A.) | At the end of the study (day 140 after the third vaccination) |
| IgG and NeutAb in the EoS | Continuous (50% titer) | SARS-CoV-2 Pseudo-virus (psSARS-2) Neutralization Assay | At the end of the study (day 140 after the third vaccination) |
| **Variables** | | | |
| Sex | Female/male | As defined in SMC' files | Current |
| Age | Continuous (years) | As defined in SMC' files | At third vaccine dose |
| BMI | Categorical: <25, 25-29.99, ≥30 | BMI was calculated by weight (kg)/(height (m))^2^ according to the HCW answer to the questionnaire. | At second vaccine dose |
| Blood pressure disease | 0/1 | According to the HCW answer to the questionnaire: defined as systolic blood pressure above 140 treated with medication | At second vaccine dose |
| Dyslipidemia | 0/1 | According to the HCW answer to the questionnaire: defined as total cholesterol above 200 or LDL cholesterol above 160 treated with medication | At second vaccine dose |
| Autoimmune disease | 0/1 | According to the HCW answer to the questionnaire: defined as known autoimmune disease treated with medication | At second vaccine dose |
| Diabetes | 0/1 | According to the HCW answer to the questionnaire: defined as HbA1C>6.5 or fasting blood sugar>126 treated with medication | At second vaccine dose |
| Heart disease | 0/1 | According to the HCW answer to the questionnaire: defined as known heart disease treated with medication | At second vaccine dose |
| Lung disease | 0/1 | According to the HCW answer to the questionnaire: defined as defined as known lung disease treated with medication | At second vaccine dose |
| Coagulation disorder | 0/1 | According to the HCW answer to the questionnaire: defined as known hemorrhage or thrombosis disease treated with medication | At second vaccine dose |
| Immunosuppressed | 0/1 | According to the HCW answer to the questionnaire: defined as organ transplantation, biologic therapy, chemotherapy, steroids, splenectomy, or HIV | At second vaccine dose |
| Allergy | 0/1 | According to the HCW answer to the questionnaire: defined as a serious allergic reaction (anaphylaxis) that required immediate treatment | During the life |
| Liver disease | 0/1 | According to the HCW answer to the questionnaire: defined as cirrhosis, hepatitis, liver cancer, metabolic disorder | At second vaccine dose |
| Kidney disease | 0/1 | According to the HCW answer to the questionnaire: defined as creatinine>1.2 or GFR<60) treated with medication | At second vaccine dose |
| Pregnancy | 0/1 | According to the HCW answer to the questionnaire: defined as confirmed pregnancy by a beta HCG blood test and ultrasound fetal heartbeats detection | At second vaccine dose |
| Specific comorbidities | Categorical: 0, 1, ≥2 | Count of comorbidities that were with significant lower antibodies titers compared to healthy people during the first 5 weeks after the first vaccine dose^3^:   - Hypertension - Diabetes - Dyslipidemia - Heart disease - Lung disease - Kidney disease - Liver disease | At second vaccine dose |

Abbreviations: NeutAb= neutralizing antibodies; EoS=end of study; IgG=Immunoglobulin G; S/CO=sample cutoff ratio; SARS-CoV-2=severe acute respiratory syndrome; BMI=Body mass index; Kg=kilogram. M=meter; HCW=health care worker; LDL=low-density lipoproteins; HIV=human immunodeficiency; GFR=Glomerular filtration rate; HbA1C=hemoglobin A1C.

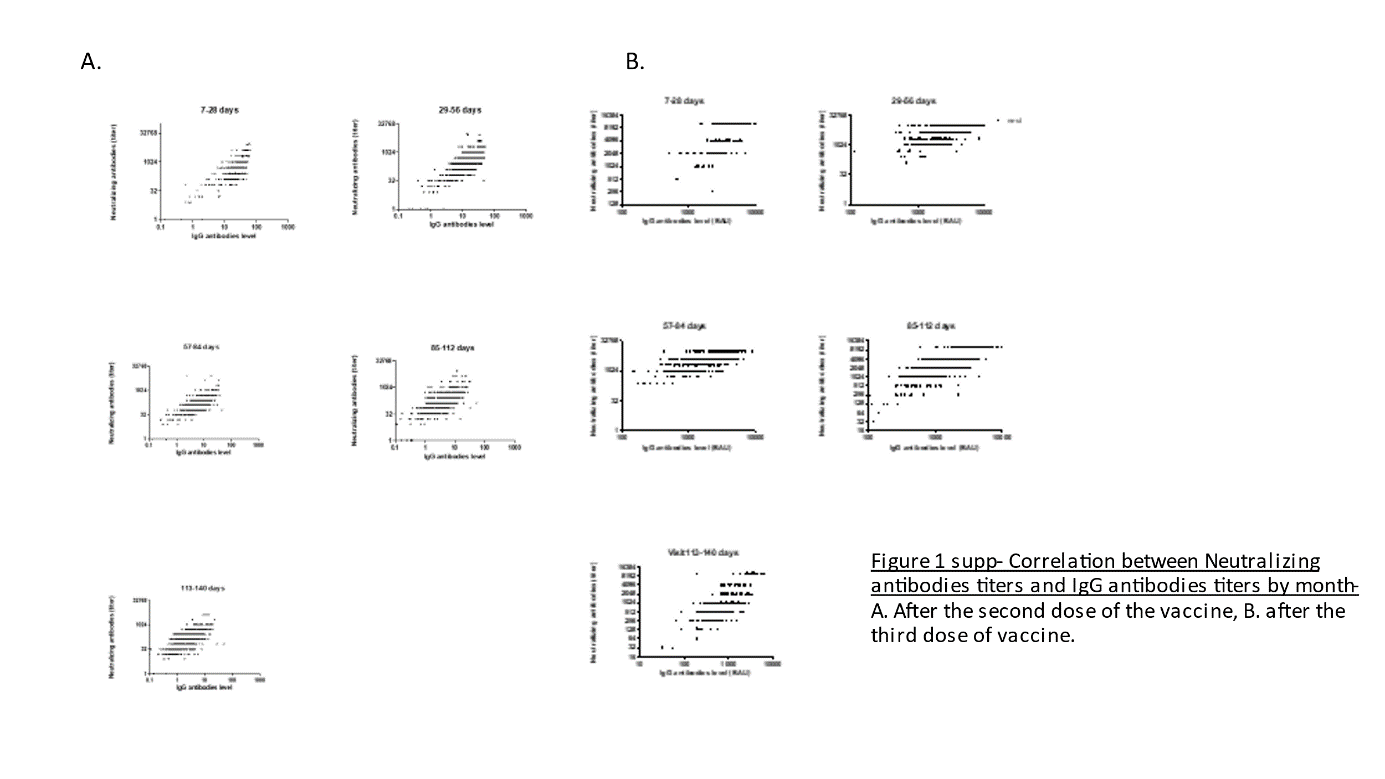

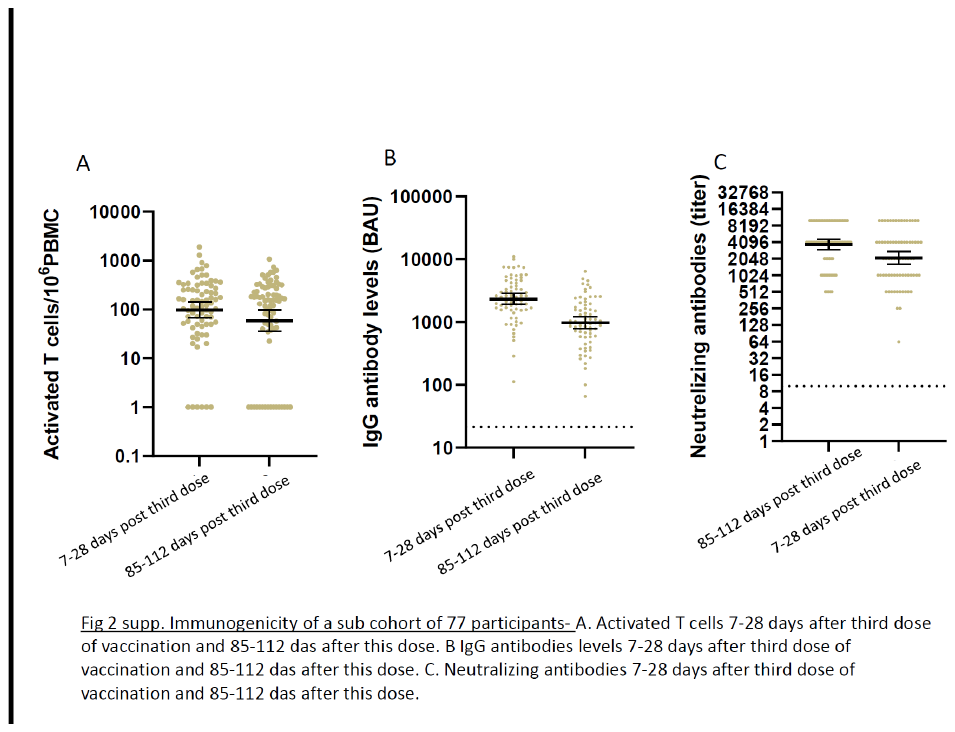
